# Randomized trial to compare acceptability of Magnesium Sulphate administration for preeclampsia and eclampsia: Springfusor pump versus standard of care

**DOI:** 10.1101/2023.05.16.23290038

**Authors:** Sam Ononge, Annettee Nakimuli, Josephat Byamugisha, Moses Adroma, Paul Kiondo, Thomas Easterling, Hillary Bracken

## Abstract

**Introduction:** In low-resource settings, magnesium sulphate (MgSO_4)_ for preeclampsia is administered majorly through an injection into the gluteal muscles 4-hourly for 24 hours. The repeated injections are very painful and may lead to infection, abscess formation and reduced compliancy.

**Objective:** To determine the acceptability of Springfusor® pump for the administration of Magnesium Sulphate in preeclampsia and eclampsia.

**Design:** Randomized Open Label Clinical Trial

**Method:** Study was conducted at Kawempe National Referral Hospital. Eligible women had systolic blood pressure of ≤40mmHg and or diastolic blood pressure >90mmHg, proteinuria ≤+1, and physician’s decision to start on MgSO_4_. Four-hundred-ninety-six participants were randomized to Springfusor® pump group or control (standard of care) administration of MgSO_4_. Intervention group had loading dose (4gm of 50% MgSO_4_ intravenously over 20 minutes) and maintenance therapy (1gm of 50% MgSO_4_ intravenously per hour for 24 hours) administered using the Springfusor®. The control group received a loading dose of 4gm of 20% MgSO_4_ IV over 15-20 minutes, followed by 10gm of 50% MgSO_4_ intramuscular (5gm in each buttock) and maintenance dose of 5gm of 50% MgSO_4_ was administered IM every 4 hours for 24 hours. Both arms received the rest of the care for preeclampsia/eclampsia as per the hospital guidelines.

Acceptability of method of administration was assessed using a Likert scale (1-5; 1 and 2: acceptable and 3-5: unacceptable). Pain at site of MgSO_4_ administration was assessed using Visual Analogue Scale 1-7, (1 minimal pain and 7 worst pain). Comparisons were assessed with X-square and Students’ t-test.

**Results:** Baseline characteristics were similar in both arms. Intervention arm was; more acceptable than the standard of care arm, (94.9% vs70.3%; p<0.001), had lower pain score (2.2±1.3 vs. 4.1±1.6; p <0.001) and fewer side effects. Maternal mortality was comparable between groups (0.8% in intervention arm vs 1.2% in the IM arm).

**TRIAL REGISTRATION:** Trial No PACTR201712002887266 (https://pactr.samrc.ac.za/)

## Introduction

Preeclampsia is a multisystem disorder that presents with a raised blood pressure and proteinuria in pregnancy [1]. Globally, preeclampsia complicates approximately 2-8% of the pregnancies [2]. The presence of convulsions with preeclampsia indicates eclampsia. Preeclampsia and eclampsia are life-threatening for both the mother and the fetus[3], and they are among the leading causes of maternal deaths and disability worldwide, especially in the low resource setting[4]. The World Health Organization (WHO) estimates that, 16% of the maternal deaths in low resource settings are due to PE/E [5]. Magnesium Sulphate (MgSO_4_) is the drug of choice for prevention and treatment of eclampsia [5]. It is administered parenterally by intravenous (IV) and or intramuscular (IM) routes. The IV therapy is commonly administered following the Zuspan regimen that requires an initial loading dose of 4 gm of magnesium sulphate over 15-20 minutesover 15-20 minutes (mins), followed by 1-2 gm hourly maintenance dose continuing for 24 hours after the loading dose or the last eclamptic fit [6]. Zuspan regimen is best delivered by electronic infusion pumps. These electronic pumps are expensive and require electricity or battery to run, making them less appropriate in low resource settings. In many low resource settings, MgSO_4_ administration follows the Pritchard regimen. The regimen is particularly complex and requires both the IV and IM administration of MgSO_4_. The loading dose of 4 gm is often delivered using an IV-push, in which clinicians slowly injects magnesium sulphate with a syringe over 15-20 mins. This is immediately followed by IM injection of 10 gm of magnesium sulphate into the gluteal muscles (5 gm on each buttock). The maintenance dose of 5 gm IM injection is administered every 4 hours for 24 hours. These repeated IM injections are painful, and can increase the risk of abscess development. Because of pain associated with the IM injection, some providers do not administer maintenance therapy and patients too may discontinue the maintenance dose for the same reason. In addition, the Pritchard regimen requires different dilutions for IV and IM doses, and different doses for loading, and maintenance doses. This regimen requires a 20% dilution of magnesium sulphate for the IV loading dose, which necessitates the health providers to calculate the quantity of sterile water to add to the magnesium sulphate solution. In most settings, health providers do not encounter eclampsia very often; and when they do, trying to remember the complex regimen is challenging.

In low resources settings that cannot afford electronic infusion pumps, there was a need to test alternative devices that can effectively and safely deliver MgSO_4_ at cheaper cost, and acceptable to the patient and health provider. The Springfusor® pump and flow control tube (FCT) designed by Go Medical Industries Pty Ltd based in Australia [7], is a promising alternative to Pritchard method of administering MgSO_4_, and is designed to simplify continuous IV infusions. The Springfusor pump does not require electricity and it is reusable. The Springfusor is powered by the potential energy stored within a spring at the heart of the device. The spring is compressed by the action of loading the Springfusor with the compatible syringe and FCT. The spring provides a constant force to the barrel of the loaded syringe. The flow rate is controlled by the FCT which offers consistent resistance to produce a steady flow. The FCT is a fine bore tube designed to provide a metered constant flow for IV infusion. FCTs has easy fitting to the patient’s cannula. The FCT exist in a variety of flow rates which enables the user to attain the desired output for exact IV delivery needs. For this study we used two varieties of FCT; the loading dose and maintenance dose. While the Springfusor can be reused indefinitely on different patients, the FCT must be replaced after each use.

The Springfusor syringe infusion pump is a low cost, non-sterile, reusable pump that requires no external power source. It is simple to use and setup, and requires only minimal training to load and operate. It is lightweight, portable and therefore does not limit the mobility of the patient.

The Springfusor has been used to administer MgSO_4_ in the treatment of severe preeclampsia in India[8] [9]. In India, Mundle and colleagues compared the manually administered IV loading dose followed by maintenance therapy given by IM route of administration via a syringe, to a loading dose and maintenance therapy given through IV infusion administered by a Springfusor device. Though there were no difference in maternal and neonatal morbidity, the Springfusor had few side effects[9]. Later, Easterling et al compared the Springfusor administration of continuous IV infusion of magnesium sulphate and 2 hourly IV boluses, the clinical findings were not different in the two groups[8]. Earlier in 1994, Freebairn et al in Australia compared the Spingfusor infusion device to intermittent boluses on administration of muscle relaxant. They were able to show that Springfusor provided a more constant level of paralysis compared to intermittent bolus administration [10]. The objective of this study was to assess the acceptability and safety of Springfusor pump for intravenous delivery of magnesium sulphate for the treatment of preeclampsia and eclampsia.

## Methods

The trial protocol is submitted as a supplementary file (S2). The study was randomized open label clinical trial conducted at Kawempe national referral and teaching hospital in Kampala district Uganda. The hospital is a government public facility and its Maternal Fetal Unit admits and delivers approximately 2100 pregnant women per month. From the facility records, 7% of the admissions are preeclamptic/eclamptic women. The women with preeclampsia and eclampsia are managed by cadres of health workers ranging from senior consultant obstetricians to medical officers, while the midwives administer the medication and nursing care.

The study included women with age of 15 years and above, with a pregnancy of 20+ weeks of gestation or had childbirth within 24 hours, presenting with preeclampsia and eclampsia i.e., have a raised blood pressure (systolic of ≤140 mmHg and/or diastolic ≤ 90mmHg), proteinuria ≤1+. We excluded women who had received MgSO_4_ 24 hours prior to admission, or had known allergy to MgSO_4_ and known elevated serum creatinine (>1.2 mg/dl) before enrolment.

Intervention: Women in the intervention arm (Springfusor® group) had the loading and maintenance therapy of MgSO_4_ using IV infusion administered using a Springfusor® infusion pump. The loading dose was 4 gm of 50% MgSO_4_ in 10 ml syringe administered over 20 mins. The infusion rate was determined by the flow control tubing calibrated to deliver 10 ml of saline over 5 mins (this system was demonstrated to deliver 4gm of 50% MgSO4 in 20 mins)[9].The maintenance dose of 4 gm of 50% MgSO_4_ in 8 ml was administered over 4 hours and infusion rate was determined by a second flow control tubing calibrated to deliver 10 ml of saline over 60 mins (this system was demonstrated to deliver 4gm of 50% MgSO4 in 4 hours)[9]. The 4-gm dose of MgSO_4_ was repeated every 4 hours for 24 hours.

Women in the control arm (standard of care) had MgSO_4_ administered according to the standard hospital practice. The loading dose of 4 gm of 20% MgSO_4_ in 20 ml syringe was administered using an IV infusion over 15-20 mins. This was immediately followed by IM injection of 10 gm of 50% MgSO_4_ mixed with 1 ml of lignocaine into the gluteal muscles (5 gm on each buttock). This was followed by maintenance dose of 5 gm IM injection, administered every 4 hourly for 24 hours.

### Study procedure

Between March and September 2019, women admitted at Kawempe hospital maternal fetal unit were screened and enrolled into the study if they meet the inclusion criteria. The study team obtained a written informed consent from the participant or from the relative if the mother was eclamptic. Those with eclampsia provided individual written consent when they regained their consciousness. Both arms received the care as per national guidelines which included management of high blood pressure using antihypertensives, laboratory investigations (urine analysis, complete blood count, renal function tests and liver function tests), prevention/treatment of fits and delivery as planned by the attending physicians. Upon enrolment, study participants were randomized to intervention or standard of care arms. The prevention and treatment of fits were started as per hospital guidelines. The time it took to administer the loading and maintenance doses in both arms were measured in mins by study nurse using a stop clock. The time was measured from the loading of the MgSO_4_ into syringe to completion of administration of the medicine in the syringe. For this paper we report the duration in mins it took to administer the loading dose and the second maintenance dose.

The study participants’ respiratory rates were monitored by the study midwives every 5 mins during the loading dose for 30 mins and hourly during the 24 hours maintenance dose administration. In addition, urine output and tendon reflexes were monitored hourly and documented in the source documents. The side effects of MgSO_4_ like nausea, vomiting, flushing of the skin, muscle weakness, confusion and drowsiness were reported 4 hourly and were captured in questionnaire. The study participants were followed till discharge from the hospital and pregnancy outcomes were extracted from the participants’ records or charts.

The primary outcome was acceptability of the method of MgSO_4_ administration assessed using Likert scale (1-5; 1: acceptable and 5: unacceptable), administered after the last maintenance dose. The method of MgSO_4_ administration was regarded acceptable if participant rated it 1 or 2, and was unacceptable if participant scored it 3 or more. The secondary outcomes were 1) the level of pain experienced during administration of MgSO_4_ and this was assessed using a visual analogue scale 1-7. The least (one) representing no pain and maximum (7) representing the worst pain imaginable, 2) proportion of complications in two arms measure as proportion of preeclamptic women who developed any one of the following; MgSO_4_ toxicity, abnormal liver and renal function test as reported by the laboratory report, an infection at the injection as reported by the clinician or a maternal deaths; 3) discontinuation rate was assessed as study participants who did not complete the recommended doses of MgSO_4_ in 24 hour period.

Participants who discontinued the method of administration by choice, due to side effects or by health workers decision were counted in each group, and; 4) reliability of Springfusor® pump in the delivery of MgSO_4_ measured as time taken by the device to deliver the MgSO_4_ as per the clinical recommendations.

### Sample size, randomisation and statistical analysis

Based on the 31% acceptability of MgSO_4_ administration using the Pritchard regimen (IM maintenance) in the Mundle et al trial[9], we calculated that we needed a sample size of 219 participants in each arm to give 90% power to detect 50% difference between the Springfusor arm and standard of care, with alpha of 0.05. With approximately 10% non-response, we needed 241 women in each arm.

Randomization was performed by a biostatistician not involved in the clinical trial who developed the allocation sequence using an online computer random number generator in block sizes of 4 and 6. The allocation sequence was concealed from the research team enrolling and screening participant in serially numbered sealed opaque envelopes (concealed allocation) containing the randomization group. After the research nurse had obtained the consent, she opened the next envelope to determine the group assignment only after the participant is enrolled, completed all the baseline assessment and it is time to allocate the intervention.

Because of the nature of the study, it was difficult to blind the implementation of the allocation and measurement of the outcome.

Descriptive statistics were used to summarize baseline characteristics of study participants and assess if randomization was successful. Acceptability and safety data were evaluated using intention-to-treat (ITT) analysis. The primary outcome (acceptability of administration of MgSO_4_ using Springfusor) was assessed using a Likert scale ranging from one (very acceptable) to five (very unacceptable). The method of MgSO_4_ administration was regarded acceptable if participant rated 1 or 2 on the Likert scale. It is not acceptable if the rating was three and above. Women who discontinue the method of administration by choice or due to side effects were considered in group of the unacceptable. The acceptability was compared in the two groups using chi-square. Secondary outcomes (complications of preeclampsia and its management and discontinuation rates) were compared using chi-square. While the level of pain was compared using means. The reliability of the Springfusor to administer MgSO_4_ in the prescribed time was assessed for the loading period and the maintenance period.

### Ethical considerations

Approval for this research was provided by the Makerere University School of Medicine Research and Ethics Committee (REC Ref 2018-015) and the Uganda National Council for Science and Technology (HS 2365). Study participants provided written informed consent. The trial was registered with Pan African Clinical Trails PACTR201712002887266. The results are reported in accordance with the CONSORT statement for randomized control trials[11] and checklist is provided as supporting file

## Results

Participants flow: Eligible participants were recruited from March to August 2019. Approximately, ten thousand (9828) women admitted at the maternal fetal unit of Kawempe national referral hospital during the study period were screened for eligibility (Fig. 1). A total of 496 eligible participants were randomized to intervention or standard of care. The loss-to-follow-up was similar in both arms. The 5 referrals to other facilities were for renal consultations to rule out possible acute kidney injury and Mulago Women Specialized Hospital to decongested Kawempe hospital when it was overloaded with patients.

**Fig. 1.**
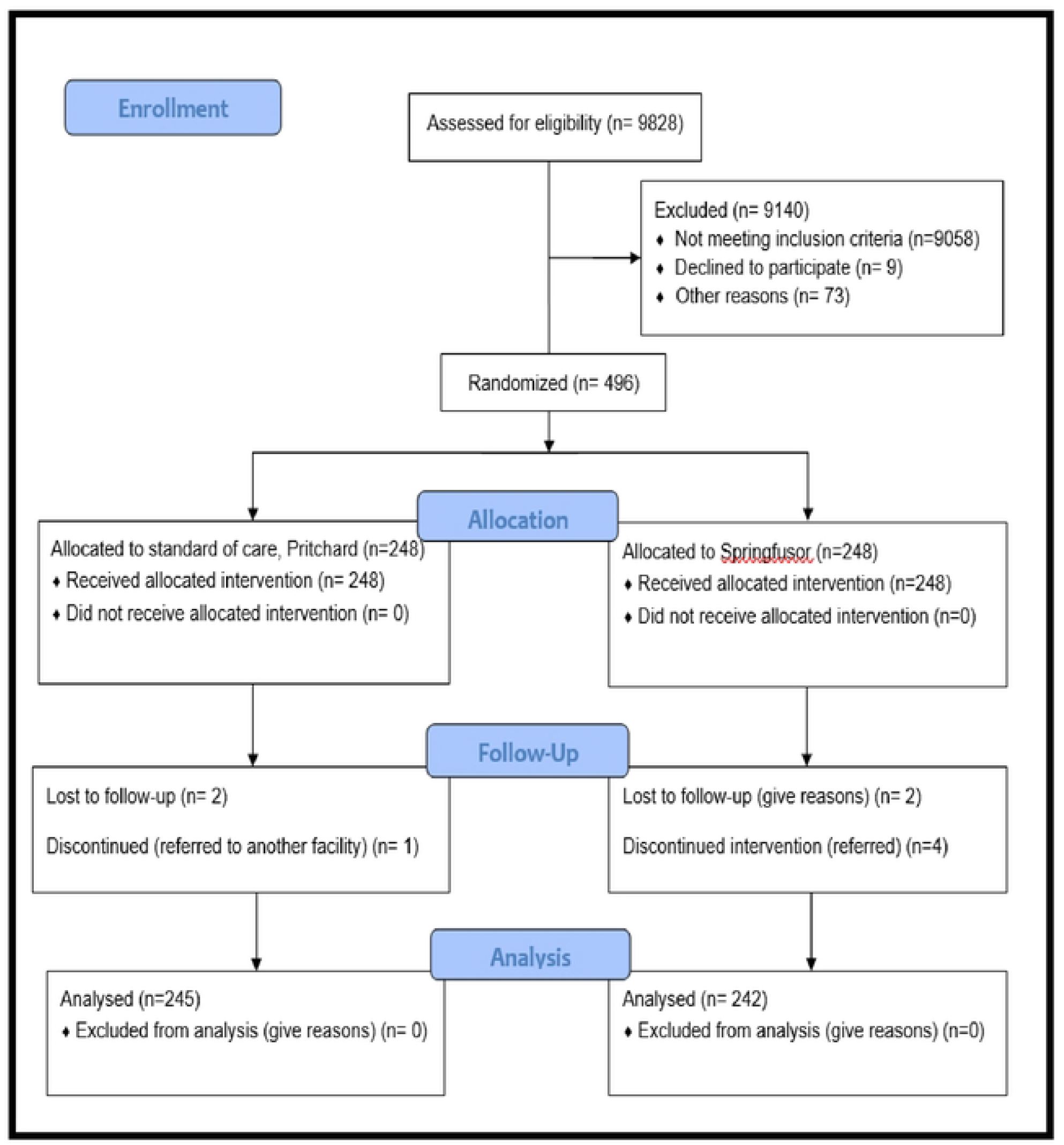
Consort diagram showing the participants flow

### Baseline characteristics

Table 1 shows the baseline characteristics of the women enrolled in the control and interventions arms. The baseline data established that randomization of the two arms were similar for almost all variables except for presenting as a postpartum and multiple pregnancy. The differences in the 2 variables were not statistically significant. More than two thirds of participants were referrals from lower facilities Table 2 shows the laboratory parameters of the study participants enrolled into the control and intervention arms. The baseline laboratory parameters were similar in the two with the exception of low level of platelets and elevated serum creatinine in the intervention arm.

**Table 1.** Demographic and clinical characteristics of the study participants

| Particulars | Standard of care<br>(Pritchard) arm<br>N=248 (%) | Springfusor arm<br>N=248 (%) |
| --- | --- | --- |
| Age in years |  |  |
| Mean (SD) | 27.2 (5.5) | 26.5 (5.6) |
| Range | 17 - 42 | 16 -45 |
| Gravidity n (%) |  |  |
| 1 | 83 (33.5) | 94 (37.9) |
| 2 | 41 (16.7) | 39 (15.7) |
| 3 | 37 (15.1) | 44 (17.7) |
| 4 | 30 (12.1) | 29 (12.0) |
| 5+ | 44 (18.0) | 35 (11.1) |
| Average gravida; mean (SD) | 2.7 (1.7) | 2.5 (1.7) |
| Enrolled postpartum (given birth)<br>n (%) | 13 (5.3) | 7 (2.8) |
| Gestation age at enrollment n (%) |  |  |
| 20-33 | 74 (29.8) | 61 (24.6) |
| 34-37 | 78 (31.5) | 94 (37.9) |
| 38+ | 79 (31.9) | 79 (31.9) |
| Unknown/missing/postpartum | 17 (6.9) | 14 (5.6) |
| Average gestation age in weeks at<br>enrolment; mean (SD) | 34.4 (4.8) | 34.8 (4.6) |
| Gestation age range (weeks) | 20 - 43 | 20 - 43 |
| Number of ANC visits (%) |  |  |
| None | 10 (4.0) | 8 (3.2) |
| 1-3 | 145 (58.5) | 142 (57.3) |
| 4+ | 93 (37.5) | 98 (39.5) |
| Average number of ANC visits;<br>mean (SD) | 3.0 (1.3) | 3.2 (1.3) |
| Multiple pregnancy* (%) | 21 (8.5) | 14 (5.8) |
| Admitted as n (%) |  |  |
| Referral in | 179 (72.2) | 172 (70.1) |
| Self-referral (Walk in) | 23 (9.3) | 34 (13.7) |
| Antenatal clinic at study site | 46 (18.5) | 42 (16.9) |
| Systolic BP at enrolment mmHg |  |  |
| Mean (SD) | 168.8 (22.4) | 167.0 (21.0) |
| Range | 126 - 237 | 130 -270 |
| ≥160 mmHg (%) | 145 (58.5) | 151 (60.9) |
| Diastolic BP at enrolment mmHg |  |  |
| Mean | 111.5 (14.6) | 112 (13.2) |
| Range | 78 - 173 | 90 -162 |
| ≥110 mmHg (%) | 129 (52.0) | 142 (57.3) |
| Enrolled as Eclamptic (%) | 14 (5.6) | 11 (4.4) |

**Table 2.** Laboratory characteristics of the participants enrolled into the study

| Particulars | Standard of care<br>N=248 (%) | Springfusor n=248 (%) |
| --- | --- | --- |
| Low platelets (<100x10<sup>3</sup>/μmoll) | 20 (8.1) | 29 (11.7) |
| Elevated alanine transaminase<br>(>31μmol/L) | 46 (18.5) | 40 (16.1) |
| Elevated aspartate transaminase<br>(>32μmol/L) | 70 (28.2) | 69 (27.8) |
| Raised bilirubin (>3.4μmol/L) | 87 (35.1) | 80 (32.2) |
| Elevated serum creatinine<br>(≥1.2mg/dL)* | 20 (8.1) | 38 (15.3) |

\*Almost all the results were available for the attending physicians after the prevention and treatment of fits with magnesium sulphate was initiated or completed. None of the participants with elevated serum creatinine was excluded from the study.

The duration of MgSO_4_ administration of loading dose was longer in the standard of care than in Springfusor arm (table 3). Few women in both groups discontinued MgSO_4_ (did not complete the recommended 24-hour doses) and the rates were similar (5.3% vs 5.0% p=0.862).

**Table 3.**
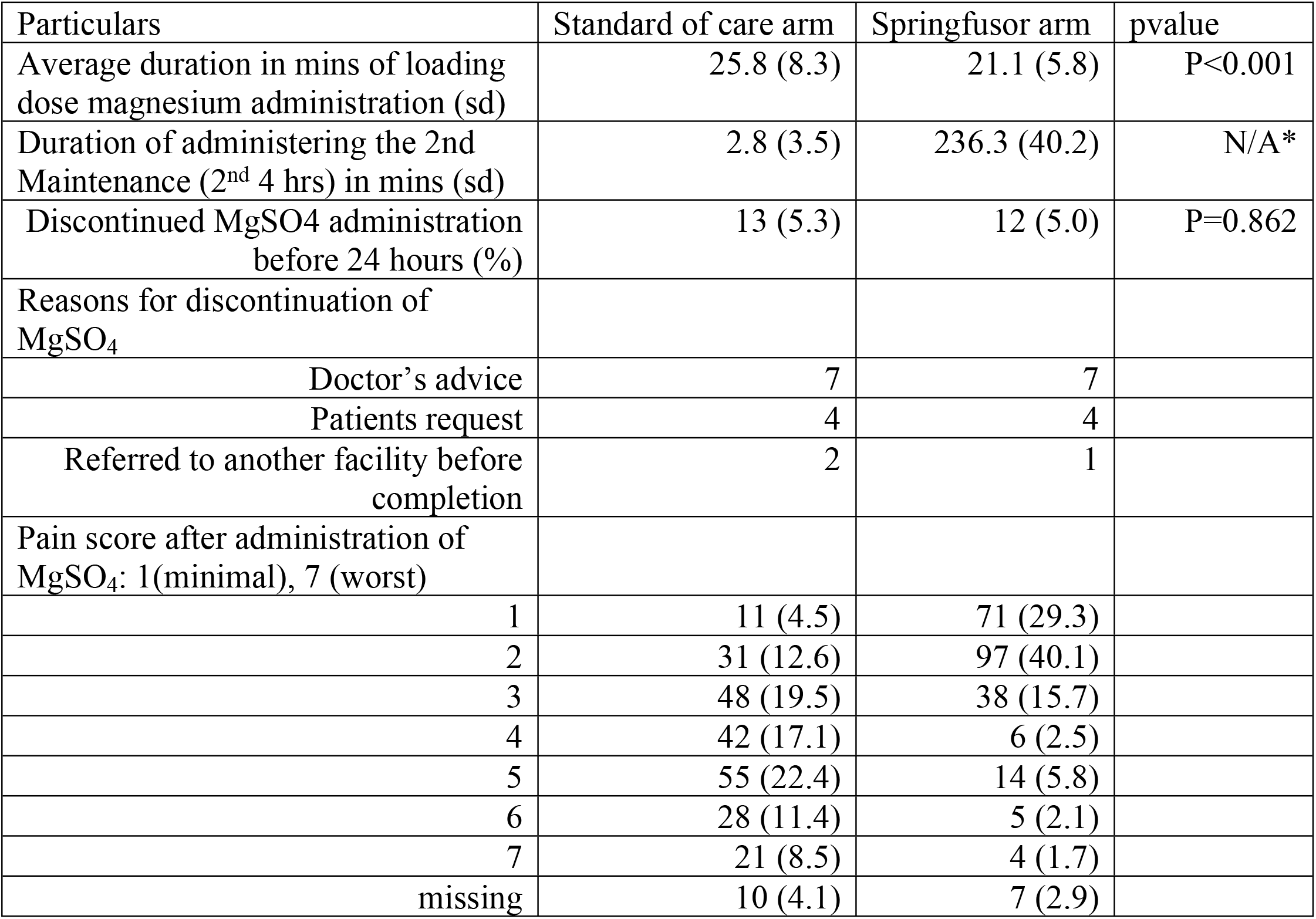
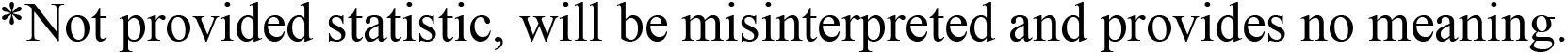
Drug administration and pain score registered by study participants

| Particulars | Standard of care arm | Springfusor arm | pvalue |
| --- | --- | --- | --- |
| Average duration in mins of loading dose magnesium administration (sd) | 25.8 (8.3) | 21.1 (5.8) | P<0.001 |
| Duration of administering the 2nd Maintenance (2<sup>nd</sup> 4 hrs) in mins (sd) | 2.8 (3.5) | 236.3 (40.2) | N/A* |
| Discontinued MgSO<sub>4</sub> administration before 24 hours (%) | 13 (5.3) | 12 (5.0) | P=0.862 |
| Reasons for discontinuation of MgSO<sub>4</sub> |  |  |  |
| Doctor's advice | 7 | 7 |  |
| Patients request | 4 | 4 |  |
| Referred to another facility before completion | 2 | 1 |  |
| Pain score after administration of MgSO<sub>4</sub>: 1(minimal), 7 (worst) |  |  |  |
| 1 | 11 (4.5) | 71 (29.3) |  |
| 2 | 31 (12.6) | 97 (40.1) |  |
| 3 | 48 (19.5) | 38 (15.7) |  |
| 4 | 42 (17.1) | 6 (2.5) |  |
| 5 | 55 (22.4) | 14 (5.8) |  |
| 6 | 28 (11.4) | 5 (2.1) |  |
| 7 | 21 (8.5) | 4 (1.7) |  |
| missing | 10 (4.1) | 7 (2.9) |  |

\*Not provided statistic, will be misinterpreted and provides no meaning.

As shown in table 4, almost all women in the intervention group found Springfusor administration of MgSO_4_ acceptable (95%) compared to the standard of care (70%) and this was a statistically significant (chi 49.7 p<0.001). The women in the standard of care arm had a higher mean pain score than the intervention arm (standard of care: 4.1(±1.6); Springfusor: 2.2(±1.3) mean difference 1.9, CI: 1.8-3.0 p<0.001). More women in the intervention arm would recommend Springfusor method of administration of MgSO_4_ to a friend compared to standard of care (96% vs 61%). Similarly, almost all women in the intervention arm would use the Springfusor for magnesium administration when they have raised blood pressure in the next pregnancy compared to control arm who would Pritchard method (96.5% vs 66%).

**Table 4.** Acceptability of MgSO_4_ administration using Springfusor compared to standard of care

| Outcome | Control group<br>(SOC)<br>N=236(%) | Intervention<br>group<br>(Springfusor)<br>N=235(%) |  | p-value |
| --- | --- | --- | --- | --- |
| **Primary outcome** |  |  |  |  |
| Acceptable ≤2 score on Likert scale (%) | 166 (70.3) | 224 (95.3) |  | <0.001 |
| **Secondary outcomes** |  |  |  |  |
| Discontinuation; Not completed the 24 hrs (%) | 13 (5.5) | 12 (5.1) |  | 0.862 |
| Mean pain score during administration (sd) | 4.1 (1.6) | 2.2 (1.3) | Mean diff<br>1.9 (CI: 1.8<br>– 3.0) | <0.001 |
| Would recommend method of administration of MgSO<sub>4</sub> to friend (%). | 145 (61.4) | 226 (96.2) |  | <0.001 |
| Would use it in future she got raised BP in next pregnancy | 155 (65.7) | 225 (95.7) |  | <0.001 |

The side effects like flushes, nausea, vomiting, drowsiness, and diplopia were more in the standard of care than in springfusor (table 5). However, the adverse events like respiratory depression, depressed patellar reflex and cardiac arrest were very few and were comparable in the two arms

**Table 5.**
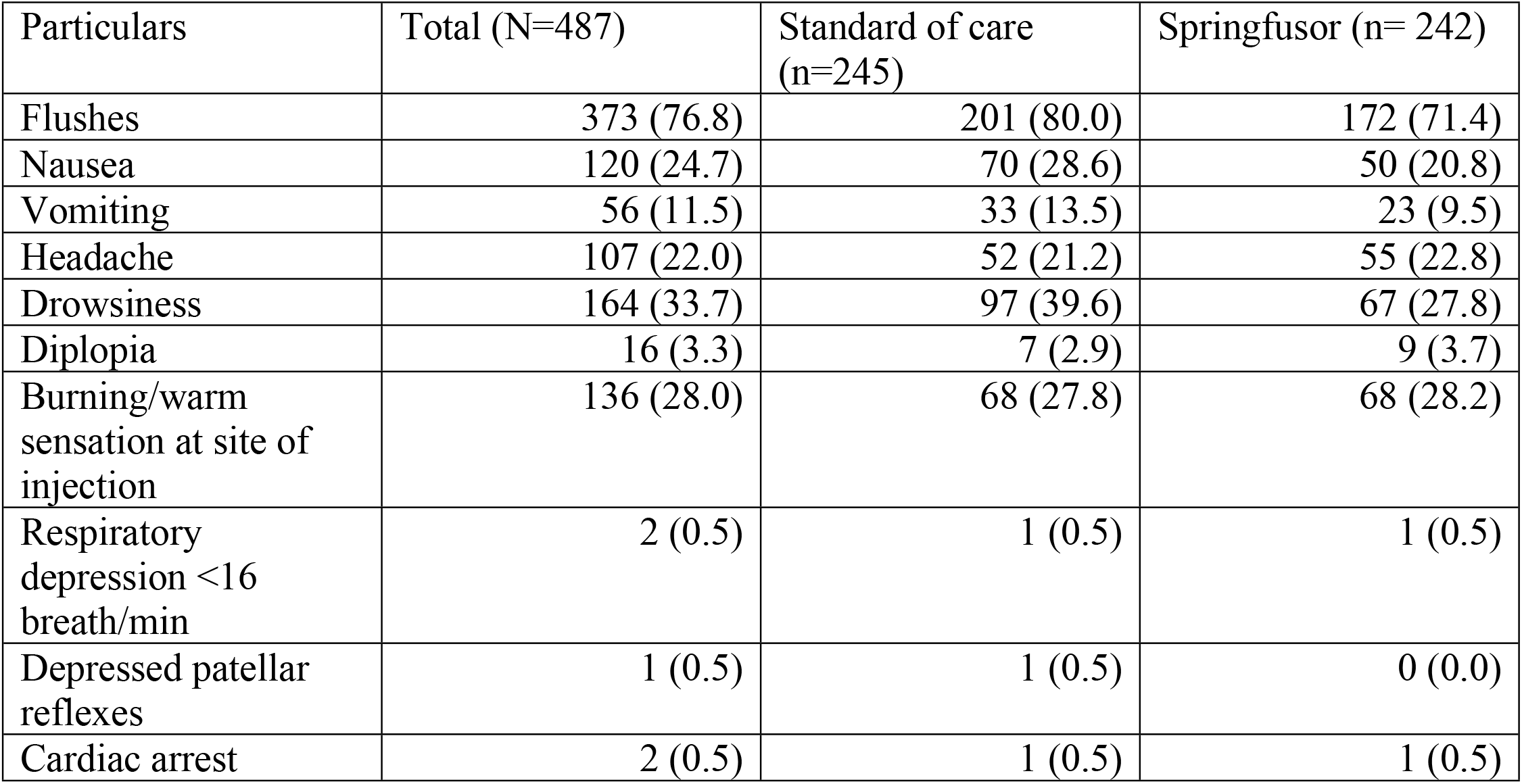
Adverse and Side effects experienced during the 24 hours of drug administration

Approximately 6% of the participants were discharged from the hospital while still pregnant after the blood pressures were controlled (table 6). Though outcomes of pregnancy were comparable in both arms, more women were delivered by caesarean section in the Springfusor arm than SOC. There were approximately 3% of the participants were referred to renal physician for specialized care (including dialysis) and follow up. Unfortunately, there were 5 maternal deaths in the whole study.

**Table 6.** Pregnancy outcome of the study participants

| Characteristics | Total (%) | Standard of care (%) | Intervention (%) |
| --- | --- | --- | --- |
| Discharged while still pregnant | 28 (5.8) | 18 (7.4) | 10 (4.1) |
| Mode of delivery (n=458) |  |  |  |
| Caesarean | 201 (43.9) | 80 (35.2) | 121 (52.4) |
| Vaginal | 257 (56.1) | 147 (64.8) | 110 (47.6) |
| Outcome of the delivery (fetus)n=488 |  |  |  |
| Alive | 398 (81.6) | 195 (80.9) | 203 (82.5) |
| Still births | 84 (17.2) | 43 (17.8) | 41 (16.7) |
| missing | 6 (1.2) | 3 (1.2) | 3 (1.2) |
| Weight of the baby |  |  |  |
| <2500 gm | 244 (50.5) | 126 (52.3) | 118 (48.8) |
| ≥2500 gm | 239 (49.5) | 115 (47.7) | 124 (51.2) |
| Referred to Nephrology hospital for further renal care n=487 | 14 (2.9) | 8 (3.3) | 6 (2.5) |
| Maternal death | 5 (1.0) | 3 (1.2) | 2 (0.8) |
| No convulsion after loading dose | 0 | 0 | 0 |

## Discussion

The definitive treatment of preeclampsia is the delivery of the fetus and placenta. However, before and after delivery of the baby and placenta, the goal of management is to control the blood pressure to normal range and minimize the development of complications like eclampsia. MgSO_4_ is the anticonvulsant for eclampsia prophylaxis and treatment[12]. MgSO_4_ therapy could be given by continuous IV infusion (Zuspan) [6] or by administering an IV bolus and IM doses for the loading dose followed by IM injections every 4 hours (Pritchard) used [13]. In low resource settings due to unavailability of the electronic pumps, the Pritchard regimen is the standard of care. The loading dose of 4gm of MgSO_4_ is often delivered via an IV-push over 15-20 mins. This process is challenging for the provider and difficult in a busy ward, and is associated with inconsistent flow rates. If administered faster than the recommended 15-20 mins, the IV-push may lead to increased pain, nausea, vomiting and flushing. Meanwhile the repeated IM injections that follow the loading dose are associated with pain, hot flushes, somnolence, and sometimes abscess formation at the site of injection.

We assessed the acceptability and safety of Springfusor pump for intravenous delivery of MgSO_4_ for prophylaxis and treatment of eclampsia among women admitted at Kawempe national referral hospital. Acceptability of intravenous administration of MgSO_4_ using Springfusor was higher compared to Pritchard regimen (standard of care). The level of acceptability of intravenous administration of MgSO_4_ using Springfusor was comparable to study by Mundle et al in India that found that it was 97%[9]. The low acceptability associated with standard of care is most likely due to pain that follows IM injection. Literature shows that the anxiety and fear associated with pain following IM injection reduces the acceptability of treatment to the patients [14, 15]. With the four-hourly frequency of IM injection of MgSO_4_ for prophylaxis and treatment of preeclampsia and eclampsia in standard of care, injection site pain is an important concern and local guidelines propose an addition of local anesthetic agent (Lignocaine) into the drug. Despite the addition of 1 ml of lignocaine into every IM injection of MgSO_4_, our study findings showed that participants on the standard of care experienced higher pain score than women who received drug through intravenous administered through the Springfusor pump.

Majority (96%) of the participants in intervention arm responded that they would recommend IV magnesium sulfate administered using Springfusor to other patients compared to standard of care (61%). In addition, almost all participants in intervention arm (97%) compared to two thirds (66%) in standard of care would use MgSO_4_ administered using Springfusor in future pregnancy if she gets preeclampsia. Literature reports that, when the safety and efficacy of two injection routes are equivalent, health care providers should consider more about patient preference because it will ensure optimal treatment adherence and ultimately improve patients experience or satisfaction[16, 17]. In our study findings showed that no women developed a fit after enrolment, indicating that both routes and dosing prevented and control the convulsions adequately.

Over all, the discontinuation rate was low (5.1%) and there was no difference in the two arms. The low rates of discontinuation could be because this assessment was among participants that participated in research study environment which has ample opportunities for questions, comments, and explanation of the process. In the real world, it is unlikely that this level of support will be available. Slightly more than half (56%) of the discontinuation were due to physician on duty recommendation. The other reasons for discontinuation were due to participants request, and three patients referred to seek treatment in another facility.

The Springfusor® pump and flow control tube (FCT) is an encouraging alternative to repeated IM administration of MgSO_4_, designed to make simpler, the continuous IV infusions. It does not require electricity and it is reusable. The Springfusor is powered by the potential energy stored within a spring at the heart of the device (Fig. 2) suitable for low resource settings. The FCT exist in a variety of flow rates which enables the user to attain the desired output for exact IV delivery needs. For this study we used two varieties of FCT as shown in the figure 2; the loading dose and maintenance dose. The Springfusor syringe infusion pump is a low-cost technology that requires only minimal training to load and operate. Being lightweight, and with a neck strap, it does not limit the mobility of the patient (Fig. 3).

**Fig. 2.**
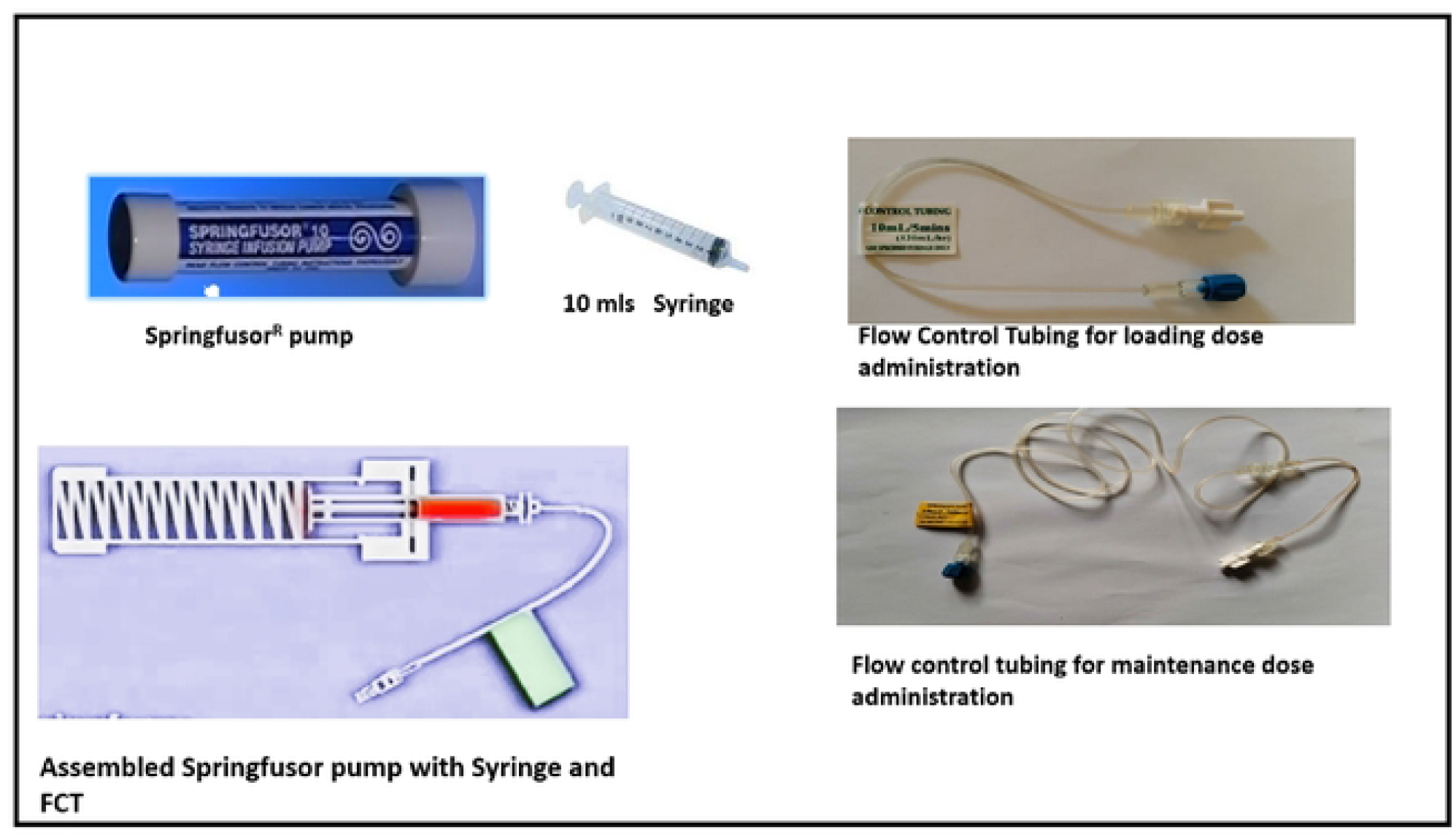
The Springfusor pump with a 10 mls syringe, flow control tubing (FCT) for administration of the loading and maintenance doses of MgSO4

**Fig. 3.**
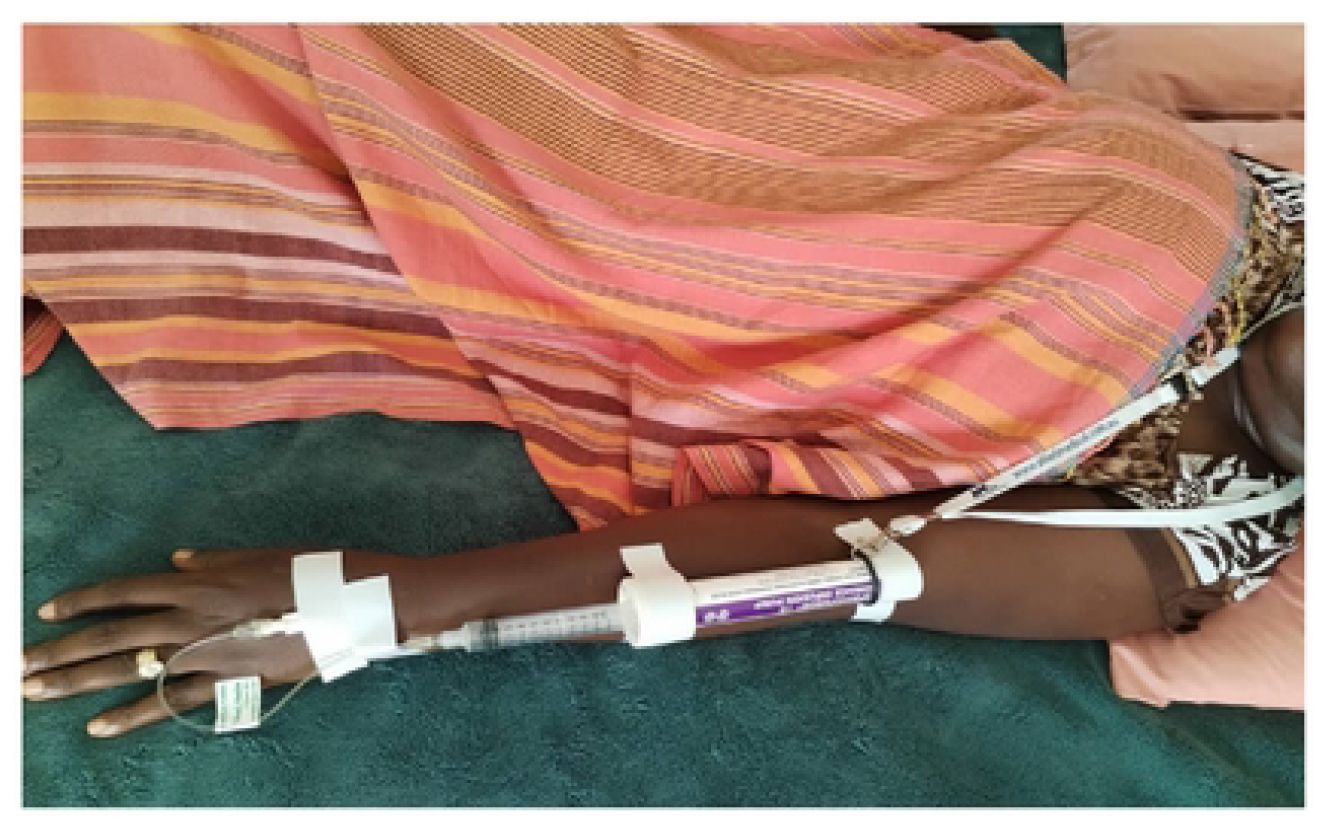
Showing participant with Springfusor pump with magnesium sulphate administered through a canula. The neck suspension string enables participants mobility without fear of dislodging the canula.

Thes study had some limitations. Firstly, the delay in accessing the laboratory work of the participants, resulted in some participants who should be excluded based on serum creatinine, enrolled into the study. The standard of care at the facility does not wait for laboratory results before starting MgSO_4_ prophylaxis. We operated within the hospital guidelines. Fortunately, no participant experienced MgSO_4_ toxicity. Secondly, we could not blind the study to the participants and providers and might have influenced the reporting of the outcomes. The nature of the study could not enable blinding of the study.

## Conclusion

Acceptance of prescribed therapy is key for adherences and to clinical outcomes, and the effect is particularly critical for treatment that require repeated injections like MgSO_4_ for preeclampsia and eclampsia. Pain associated with intramuscular injection (standard of care) was less with the intravenous infusion (Springfusor®) than with intramuscular administration. In addition, the intravenous administration was preferred to standard of care with women endorsing a greater likelihood to use it in next pregnancy or recommend it to a friend.

## Data Availability

As supplementary information in attachment

## Acknowledgement

The authors are grateful to Kawempe National Referral Hospital administration and staff for support in study implementation, data collectors for the great work done and the participants who gave in their time to participate in the study.

